# Reliability testing of patient-reported measurement instruments of Shared Decision-Making within surgical treatment pathways: a mixed-methods study

**DOI:** 10.1101/2025.06.22.25330089

**Authors:** Vignesh Gopalan, Christin Hoffmann, Hilary L Bekker, Samuel Lawday, Andy Judge, Della Hopkins, Jane Blazeby, Kerry Avery, Angus G.K. McNair

**Author notes:** **CORRESPONDING AUTHOR**, Vignesh Gopalan, Address: National Institute for Health and Care Research Bristol Biomedical Research Centre, Bristol Centre for Surgical Research, Canynge Hall (Bristol Medical School: Population Health Sciences, Bristol, BS8 2PS, UK.

## Abstract

**Objectives:** To evaluate the test-retest reliability of two instruments measuring shared decision-making (SDM) and to explore factors affecting the stability of participant’s lived experience of SDM during the test-retest interval.

**Design:** Mixed-methods prospective cohort study, nested within an ongoing National Health Service (NHS, United Kingdom) Trust Quality Improvement study

**Setting:** Seven surgical specialties within a tertiary NHS Hospital Trust.

**Participants/Procedures:** Patients (>18 years) booked for elective surgical interventions were invited to complete two measurement instruments at each timepoint: CollaboRATE (a 3-item questionnaire developed to assess patients’ perception of professionals SDM skills) and SHARED (a 10-item questionnaire to assess patients’ experience of making a treatment decision with health professionals). Instruments were completed at baseline timepoint (date of surgery booking consultation) and 5-10 days after consultation, but prior to surgery (retest timepoint).

**Main outcome measures:** Intra-class correlation (two-way, mixed, absolute agreement) and Kappa coefficients at item, total- and top-score levels were calculated. Bland-Altman plots were used to describe agreement between initial and retest measurement for each instrument.

Semi-structured interviews to explore participants’ lived experience were conducted remotely after retest measurement with a purposively selected sample of patients of varying socio-demographic characteristics, surgical specialties, and direction of score change. Transcriptions underwent thematic analysis using inductive coding approaches to identify themes.

**Results:** 86 patients completed retest measurements (median time to completion = 8 days). Test-retest reliability was weak for CollaboRATE (ICC=0.34, p<0.001) and moderate for SHARED (ICC=0.52, p<0.001) total scores. Test-retest reliability was moderate for CollaboRATE top score (κ = 0.47, p<0.001). Interviews with nine patients identified two key themes driving instability in the test-retest interval: 1) ongoing reflection on the SDM process, and 2) a need for more support for SDM.

**Conclusion:** Our study demonstrated weak-moderate reliability in measuring patient-reported SDM which may be explained by patients’ continued reflection on decisions after surgical consultations. Future research should consider the fact that SDM is a process, and work is needed to understand how and when to optimally measure SDM so that the impact of continued reflection and reasoning are not missed.

**KEY MESSAGES:** *What is already known on this topic?:*

- Instruments (e.g. self-reported questionnaires) measuring SDM from the patient perspective are widely used in clinical pathways, in research, and to guide the development of healthcare services. Few studies have evaluated the test-retest reliability of these instruments, and those done to date report poor reliability.

*What this study adds?:*

- This mixed-methods study explored the test-retest reliability of two SDM measurement instruments in the context of decision-making around surgical interventions. It is the first to supplement standard test-retest methods with interviews to explore patients’ perceptions of SDM over time in this setting. We found that patients reflect on decision-making processes and consider how this may affect the interpretation of reliability metrics in the context of SDM measurement.
- Our results affirm the dynamic nature of SDM and suggest that test-retest reliability may therefore not be an indicator of the overall quality of an SDM measure when used in this context.

*How this study might affect research, practice or policy – *summarise the implications of this study*:*

- SDM measurement instruments have different levels of reliability. Routine SDM measurement may identify a subgroup of patients who reflectively question their decisions for medical treatment in the time following consultation. There is scope to target interventions to improve SDM in the period between consultation and procedure. Clinicians and policymakers should design pathways with the timing of SDM measurement kept in mind.

## INTRODUCTION

Shared decision-making (SDM) is the process by which patients and clinicians work together to make decisions centred around patient values and preferences [1]. High-quality SDM leads to enhanced clinician-patient reasoning and may reduce over-treatment and health service utilisation [2–6]. A key challenge for health services is to measure the patient experience of SDM within the routine care pathway [7,8]. Appropriate measurement of SDM is critical for using evidence to improve patient experiences and delivery of care and is a key research priority for NICE [9][10].

There is little consensus on how best to evaluate SDM in the healthcare setting [11]. This is, in part, due to the large number of measurement instruments which have been created to capture the many facets of the SDM process, including the perspectives of the different people involved in decision-making, the varying clinical settings the instrument is used in, and the different goals of SDM measurement at an individual, group and institutional level [12,13].

There is a challenge in identifying SDM measurement instruments that are psychometrically robust and meaningful for use in health service research, audit and quality improvement. Test-retest reliability is the ability to provide consistent scores on a measurement instrument over time, assuming no changes have occurred in the underlying construct (i.e. a patients’ experience of SDM remain stable if no events or factors have influenced their perceptions of SDM) [14]. When measuring SDM in care pathways, it is pragmatically assumed that patients’ perceptions of SDM are ‘stable’ in between the initial consultation and subsequent healthcare intervention, as long as there are not additional points of contact with the medical team. However, few instruments measuring SDM have evidence of formal reliability testing that would support this assumption-a recent systematic review that appraised psychometric evidence for 40 SDM instruments reported only four studies evaluating test-retest reliability, all of which had a ‘poor’ methodological quality rating [11]. There is also a lack of qualitative evaluation of patient experiences alongside test-retest reliability measurement meaning that factors which influence how patients look back on decision processes have not been explored [11].

A further challenge specific to instruments measuring SDM is in assessing if scores (or score changes) are an indicator of the quality of the measurement instrument, or rather an indicator of a change in a patients’ perception of the SDM process. Instruments ask patients to reflect on their consultations, signposting to the concept of SDM. Over time, patients may keep thinking about their healthcare decision, seek more information, or talk with others about their experience. Patient perceived rating of the same experience may therefore change [15,16]. The SDM measurement instrument may have performed consistently but, as the perceived experience of SDM has changed, retest scores may consequently increase or reduce. This dynamic nature of reasoning can compromise the interpretation of scores and create challenges for clinicians, researchers and policymakers in terms of when to measure SDM, how to interpret results of SDM measurement scores and how best to integrate SDM measurement within clinical pathways.

As part of a quality improvement (QI) programme to integrate self-reported measurement of SDM for use in NHS care [8], this study used mixed methods to explore the test-retest reliability, estimate the standard error of measurement (SEM) and the minimal detectable change (MDC) of two instruments developed to assess the patient experience of SDM in healthcare: CollaboRATE [17] and the SHARED questionnaire [18]. To date, no test-retest reliability data has been reported for the either instrument. This study is also the first to use qualitative methods to explore factors affecting the stability of participants’ lived experience during the test-retest interval.

## METHODS

This study employed a mixed-methods design to assess the psychometric properties of the CollaboRATE and the SHARED instruments and the stability of patients lived experience between test and retest timepoints. The COnsensus-based Standards for selection of health status Measurement Instruments (COSMIN) handbook and COnsolidated criteria for REporting Qualitative studies (COREQ) checklist informed the study methods.

### Setting and context

We invited patients who completed SDM measurement as part of a larger QI project at a tertiary UK NHS hospital to participate in this study [8]. The purpose of SDM measurement in this QI programme was to evaluate patients’ experience of surgical SDM and use real-time feedback to change patients’ and healthcare professionals’ decision-making processes before elective surgery. Both the CollaboRATE and the SHARED SDM measurement instruments were operationalised into an online survey; patients booked for elective surgical interventions were automatically sent this survey after their appointment with a surgeon. Completion of this survey formed the baseline measurement scores for this study. The published protocol and usability analysis describes the process in further detail [8,19].

### Measurement Instruments

Both the CollaboRATE and the SHARED instruments are self-report questionnaires (SRQ) used to measure SDM. Methods of scoring each instrument utilised within this study are summarised in **Table 1**. In brief:

**Table 1.** Scoring of CollaboRATE and SHARED. (as outlined by instrument developers [17,18])

|  |  |  |
| --- | --- | --- |
|  | **CollaboRATE (3 item, 10-point scale)** | **SHARED (10 item, 5-point scale)** |
| **Continuous scores** | CollaboRATE total score<br>Raw score range: 0 – 27 (linear transformation for analysis: adjusted range 0 – 100)<br>CollaboRATE item level raw score<br>Range: 0 – 9 | SHARED ‘SDM Present’ total score<br>Score of 4 or 5 at item level = SDM present (coded as ‘1’)<br>Score of 1, 2 or 3 at item level = SDM absent (coded as ‘0’)<br>Overall range: 0 – 10<br>SHARED item level raw score<br>Range: 1 - 5<br>SHARED raw total score<br>Range: 10 - 50 |
| **Dichotomous scores** | CollaboRATE top score ‘Yes’ vs ‘No’<br>Score of ‘9’ on all 3 items (Yes) or any other score less than this (No) | SHARED item level ‘SDM Present’ vs ‘SDM Absent’<br>Scored 1 (SDM Present) or 0 (SDM Absent) |

The CollaboRATE questionnaire[17] is a ‘fast and frugal’ SRQ which demonstrates acceptable discriminative validity, concurrent validity and intra-rater reliability [11,20]. It was developed to assess patient perception of health professionals’ communication in consultations as an indicator of service provider skills to enhance the SDM process [21,22].

The SHARED questionnaire [18] is a theoretically informed SRQ assessing patient perceptions of i) professionals’ communication about the decision, ii) patient involvement in the decision and iii) the between patient-professional agreement with the choice made. It was developed for use within a NHS QI programme, has good content validity, is integrated well into care pathways, and is responsive to SDM initiatives [18,23,24].

### Participants

We invited adult (≥18 years old) patients to participate in this study if they had completed SDM measurement at the time of booking for elective surgery or an invasive surgical procedure. SDM measurement scores at this timepoint were used as ‘baseline’ scores for this study. Patients were booked for elective orthopaedic, urology, breast, vascular, neurosurgical, gastrointestinal or gynaecological surgery/procedures. We recruited patients to complete retest SDM measurement and participate in interviews between 8^th^ November 2023 and 31^st^ March 2024.

We excluded patients from this study if they were unable to give consent or refused to give consent to participate, if they were booked for unplanned or emergency surgery, or if they had already undergone the planned surgery at the time of being approached for participation.

### Data collection

Socio-economic and demographic data were collected from electronic patient records. These included age, sex, ethnicity, surgical specialty and number of appointments attended within the retest interval.

#### Procedure for retest SDM measurement

Participants were invited to complete baseline measurement of SDM through the CollaboRATE and the SHARED SRQ on day 0 (the day of initial consultation and booking for surgery) via an automated electronic survey within the QI programme [8,19]. Purposive sampling of participants completing ‘baseline’ measurements was undertaken to complete retest measurement aiming to achieve a patient sample comprising of a range of different surgical specialties and socio-demographic characteristics (age, gender, ethnicity). We aimed to achieve a sample of 50 patients completing retest measurement to meet a minimum rating of ‘good’ according to the COSMIN standard for reliability testing [25].

Eligible patients received a phone call from the study team between day 1 and day 4 after consultation (and completion of baseline SDM measurement) to provide verbal information and an invitation to participate in retest measurement as part of this study. Patients were subsequently also sent an email containing study information materials, an electronic consent form and a link to complete retest SRQ. For the purpose of retest SDM measurement the CollaboRATE and the SHARED SRQ were operationalised into an electronic survey and administered by a third-party survey site (JISC, www.onlinesurveys.ac.uk). The platform, layout and wording of the email invitation and survey did differ to baseline SDM measurement procedures as it was not possible to carry out retest measurement as part of the same automated platform used in the QI programme. Wording and response options for both SRQ were, however, identical to those presented to patients at baseline SDM measurement. Participants were instructed to complete the retest measurement within a five-day response window, between day 5 and day 10 after initial consultation, and prior to the planned surgical intervention. A minimum of five days were chosen to allow sufficient washout between tests, whilst maintaining assumed stability of patients’ experiences and minimising potential recall bias. A single email reminder was sent on day 9. Measurements were independent - at the second administration the patient was not made aware of their baseline scores.

In line with COSMIN guidelines, we endeavoured to ensure that both baseline and retest measurements were completed under the same conditions (same SRQ items, digital administration of instruments, same response options). There was no stipulation as to the setting in which the SRQ were completed (at hospital, at home etc.) at either baseline or retest timepoint. No intentional external intervention to the construct to be measured (i.e. SDM) was made within the retest interval. Patients followed the usual clinical pathways and processes outside of involvement in this study.

#### Procedure for patient interviews

A sub-set of patients who completed retest measurement were invited by phone call to participate in semi-structured interviews to explore their lived experience and factors affecting stability in the test-retest interval. The final sample size was determined when no new insights were identified from the data and sufficient data were collected to address the research question. Purposive sampling characteristics again included surgical specialty and socio-demographic characteristics but also focussed on direction (both improvement and deterioration) and magnitude of change in retest scores.

Semi-structured interviews were carried out by one member of the study team (VG) with the support of supervising members of the study team (CH, AM) who have extensive training and experience to a post-doctoral level in qualitative methodology. The interviewer had no relationship with participants prior to the commencement of the study. Interviews were carried out within four weeks of patients completing retest measurement to minimise recall bias.

A topic guide (**Appendix 1**) was developed and piloted with input from the multidisciplinary research team. Interviews focussed on eliciting reasons for changes between baseline and retest perceptions of SDM and explore the concept of ‘stability’. Interviews were conducted via phone and were audio-recorded. Notes were taken during the interviews to assist with future interviews and subsequent data analysis. Recordings were transcribed verbatim by a professional transcription service, with identifiable information anonymised during this process. Transcripts were checked for accuracy against audio recordings by one researcher (VG) prior to analysis.

### Data analysis

#### Procedure for quantitative data

Socio-economic and demographic data were analysed using descriptive statistics.

To assess reliability between baseline and retest measurement intra-class correlation coefficients (ICC; two-way, mixed effects model, absolute agreement) were calculated for continuous scores and Cohen’s kappa (κ) was calculated for dichotomous scores (**Table 1**). Values were interpreted using established cut-offs for strength of agreement [26]: Excellent >0.8; Good 0.61 – 0.8; Moderate 0.41 – 0.6; Fair 0.21 – 0.4; Poor ≤0.2. Bland-Altman plots for each measure were used to illustrate the degree of agreement [27]. Each participant is represented on the graph by plotting the mean value of the two assessments (x-axis) and the difference between the assessments (y-axis). Mean difference and 95% Limits of Agreement (LoA) were calculated.

The SEM (range within which a person’s true score may fall) was estimated using methods of within-subject standard deviation (SD). SEM is estimated using the equation ***SD x*** √***(1 − test–retest reliability)***. Subsequently, a distribution-based method was used to calculate the MDC (the lowest change in score that can be considered beyond the measurement error of the instrument, although it may not indicate clinical significance). MDC was calculated by the equation ***SEM x*** √***2 x z***. √*2* accounts for the error introduced on the two occasions the measure was completed. The *z* value represents the desired confidence level, for which we used 1.96 (95% confidence level) [28]. Ceiling (% frequency of highest possible score) and floor (% frequency of lowest possible score) effects were calculated. Floor and ceiling effects were classified as significant if ≥15% and negligible if <5% [29]. Data were managed using STATA version 17.

#### Procedure for qualitative data

Principles of inductive thematic analysis [30] were applied to transcripts, namely i) reading and re-reading transcripts and notes, ii) iterative inductive generation of codes (assigned to relevant excerpts in the data), iii) identifying initial themes by collating similar codes, iv) checking themes against the whole dataset to ensure consistency and sufficient support for the theme, v) detailed analysis linking themes across transcripts. This process was carried out until data sufficiently addressed the research question. Analyses were carried out in parallel to data collection to determine data saturation and make alterations to the interview topic guide in order to explore emerging and important topics further.

Coding and analyses were undertaken for all transcripts by VG, with a randoms subset (30%) of transcripts undergoing independent dual coding (by SL). Preliminary findings, and any uncertainties/discrepancies with regards to assigned codes were discussed between both coders. Input from the multi-disciplinary study team across several meetings after initial identification of codes and themes guided the reorganisation and focussing of themes to better answer the research question and reflect the data. Transcripts were managed with the qualitative data management software (NVivo).

### Patient and Public Involvement

We invited a patient and public contributor with lived experience of surgery who was part of the wider QI project steering group to be involved in the design and conduct of this research. Input was sought from the contributor as appropriate throughout the study (e.g. review of patient-facing materials such as survey invitation/instructions). The interview topic guide was reviewed and piloted with members of the public who matched the inclusion criteria for the study (but were not participants). Our results will be disseminated in plain English through meetings with a Patient Advisory Group involved in the QI project [8].

## RESULTS

151 patients were invited to participate in this study. Of these, 86 (57%) completed the SRQ within the retest window (i.e. day 5 – 10). All participants completed baseline and retest measurement fully, leaving no missing data. Participant characteristics are summarised in **Table 2**.

**Table 2.**
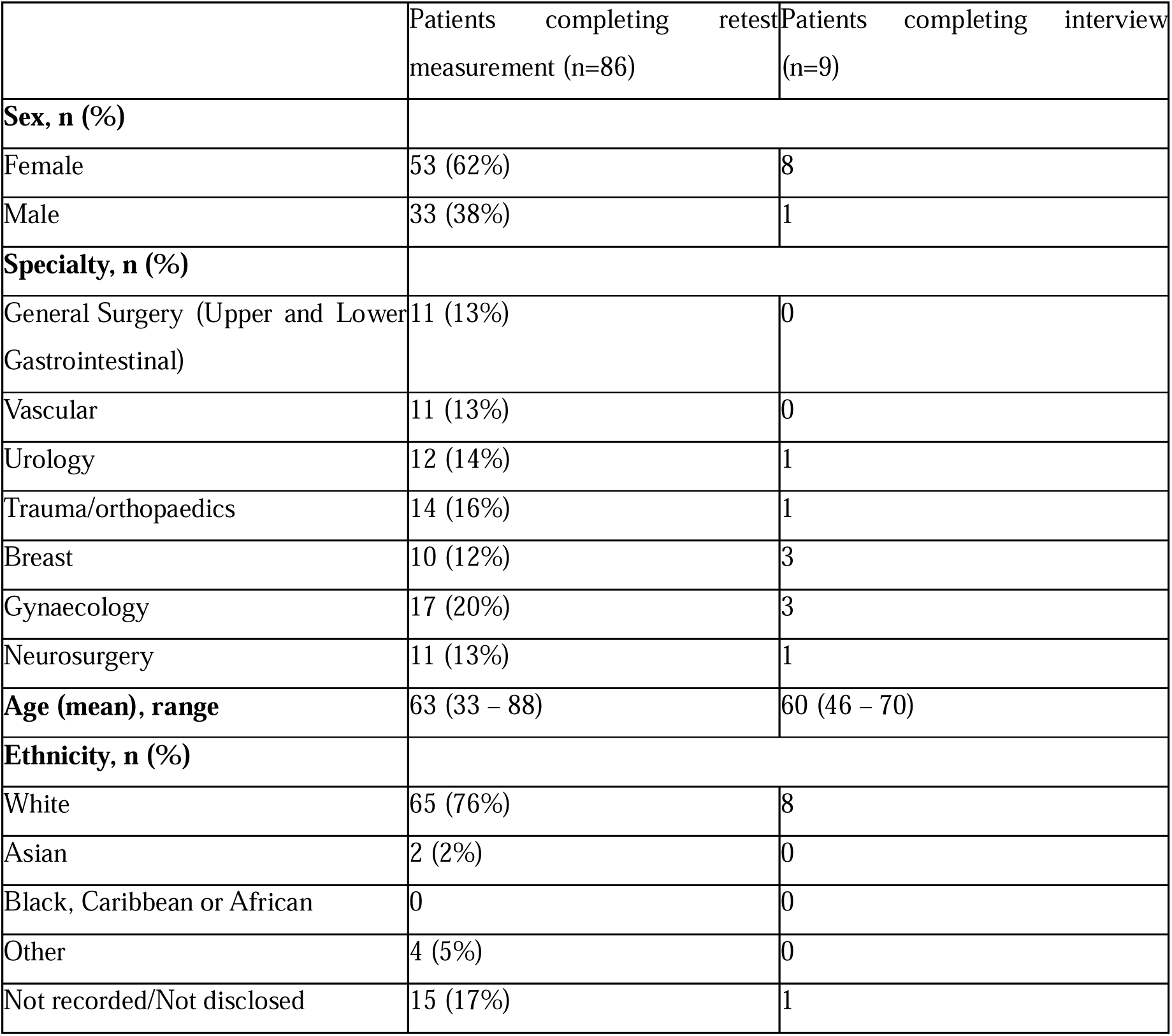
Participant characteristics.

|  | Patients completing retest measurement (n=86) | Patients completing interview (n=9) |
| --- | --- | --- |
| **Sex, n (%)** |  |  |
| Female | 53 (62%) | 8 |
| Male | 33 (38%) | 1 |
| **Specialty, n (%)** |  |  |
| General Surgery (Upper and Lower Gastrointestinal) | 11 (13%) | 0 |
| Vascular | 11 (13%) | 0 |
| Urology | 12 (14%) | 1 |
| Trauma/orthopaedics | 14 (16%) | 1 |
| Breast | 10 (12%) | 3 |
| Gynaecology | 17 (20%) | 3 |
| Neurosurgery | 11 (13%) | 1 |
| **Age (mean), range** | 63 (33 – 88) | 60 (46 – 70) |
| **Ethnicity, n (%)** |  |  |
| White | 65 (76%) | 8 |
| Asian | 2 (2%) | 0 |
| Black, Caribbean or African | 0 | 0 |
| Other | 4 (5%) | 0 |
| Not recorded/Not disclosed | 15 (17%) | 1 |

### Demographics

53 participants (62%) were female. Mean participant age was 62.9 years (range 33 – 88 years). Median timepoint of retest measurement was 7.7 days (range 5 – 10 days) after baseline measurement.

### Evaluation of test-retest reliability, SEM and MDC

#### CollaboRATE

Mean CollaboRATE total score (scaled to 0 – 100) was 97.3 (SD 0.8) at baseline and 96.1 (SD 1.12) at retest. **Table 3** summarises test-retest reliability for CollaboRATE. At item level, CollaboRATE demonstrated an ICC (2,2) of 0.268 (p = 0.006), 0.349 (p = 0.000) and 0.263 (p = 0.007) for items 1 – 3 respectively, indicating ‘fair’ levels of agreement between the two measurement points.

**Table 3.**
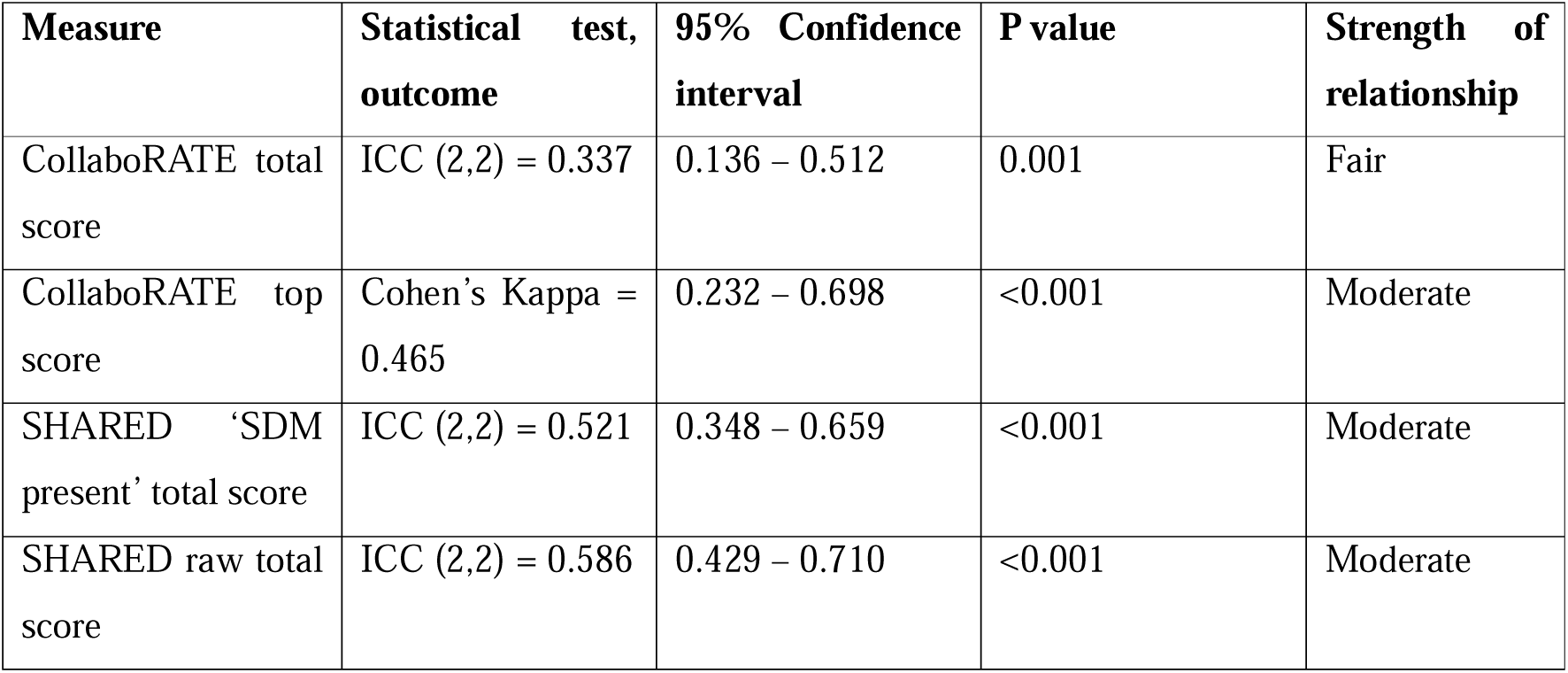
Test-retest reliability for CollaboRATE and SHARED instruments.

| Measure | Statistical test, outcome | 95% Confidence interval | P value | Strength of relationship |
| --- | --- | --- | --- | --- |
| CollaboRATE total score | ICC (2,2) = 0.337 | 0.136 – 0.512 | 0.001 | Fair |
| CollaboRATE top score | Cohen’s Kappa = 0.465 | 0.232 – 0.698 | <0.001 | Moderate |
| SHARED ‘SDM present’ total score | ICC (2,2) = 0.521 | 0.348 – 0.659 | <0.001 | Moderate |
| SHARED raw total score | ICC (2,2) = 0.586 | 0.429 – 0.710 | <0.001 | Moderate |

SEM for CollaboRATE total score was 1.99 and MDC was +/- 5.52. There were significant ceiling effects at baseline (71%) and retest (66%), with no patients giving the lowest score possible (i.e. no measured floor effect).

#### SHARED

Mean SHARED ‘SDM present’ score (i.e. a score of 4 (agree) or 5 (strongly agree) on each of 10 items, creating a possible range 0 – 10) was 8.83 (SD 0.22) at baseline and 8.59 (SD 0.25) at retest. For SHARED ‘SDM present’ total score, ICC was 0.521. For SHARED raw total score ICC was 0.586 indicating moderate strength of agreement for both methods of scoring (**Table 3**). **Table 4** summarises scores at item level.

**Table 4.**
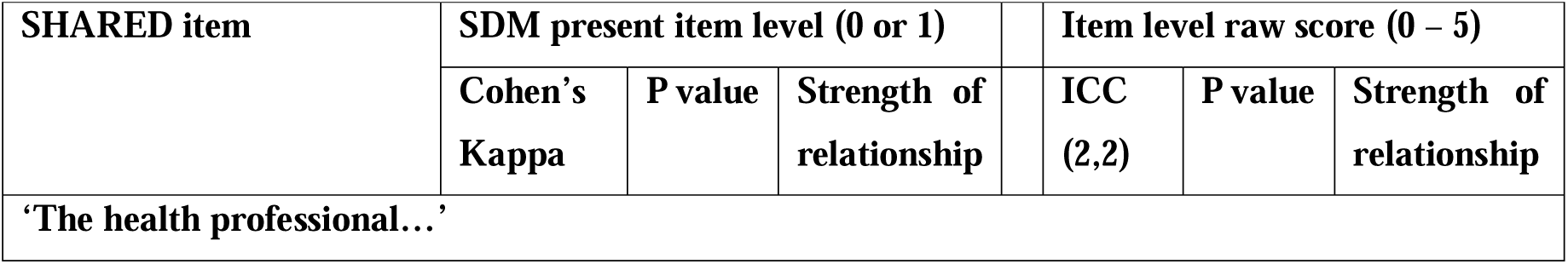
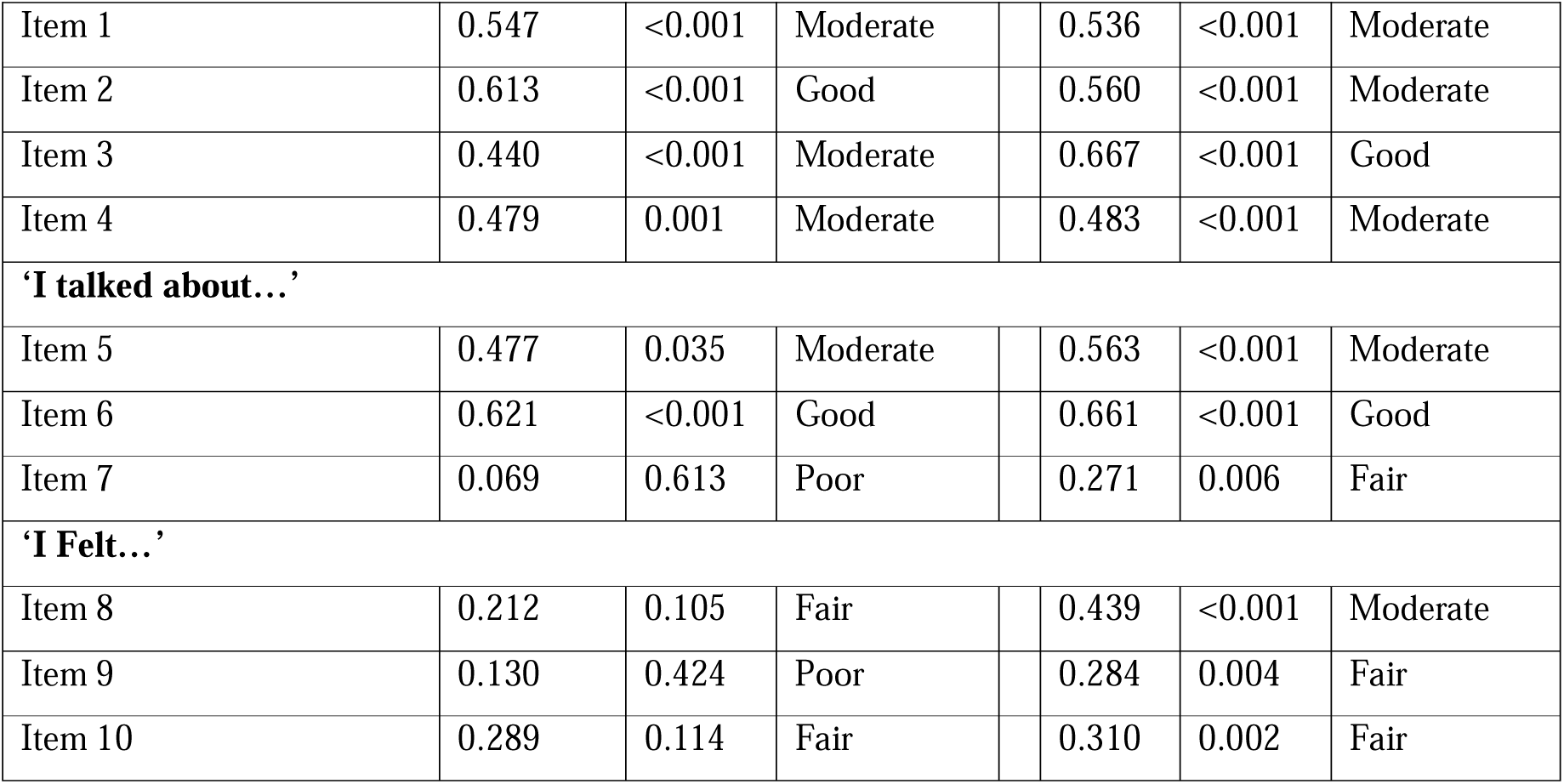
Test-retest reliability of SHARED at item level, with items grouped by how they are presented to patients completing the SRQ.

SEM for SHARED ‘SDM Present’ total score was 1.49 and MDC was +/-4.12. There were comparably significant ceiling effects at baseline (63.9%) and retest (61.6%). Floor effect at both timepoints was negligible (1.16%).

#### Strength of agreement

For CollaboRATE total score, mean difference was 1.16 (SD = 10.4, LoA range -19.3 – 21.6). For SHARED ‘SDM Present’ total score, mean difference was 0.23 (SD 2.10, LoA range -3.89 – 4.35). Bland-Altman plots for CollaboRATE (**Figure 1A**) and SHARED (**Figure 1B**) demonstrate no clear systemic bias. However, both instruments have wide LoA and outlines which may indicate instances of high variability in perceived SDM across timepoints. Qualitative interviewing explored this in further detail.

**Figure 1.**
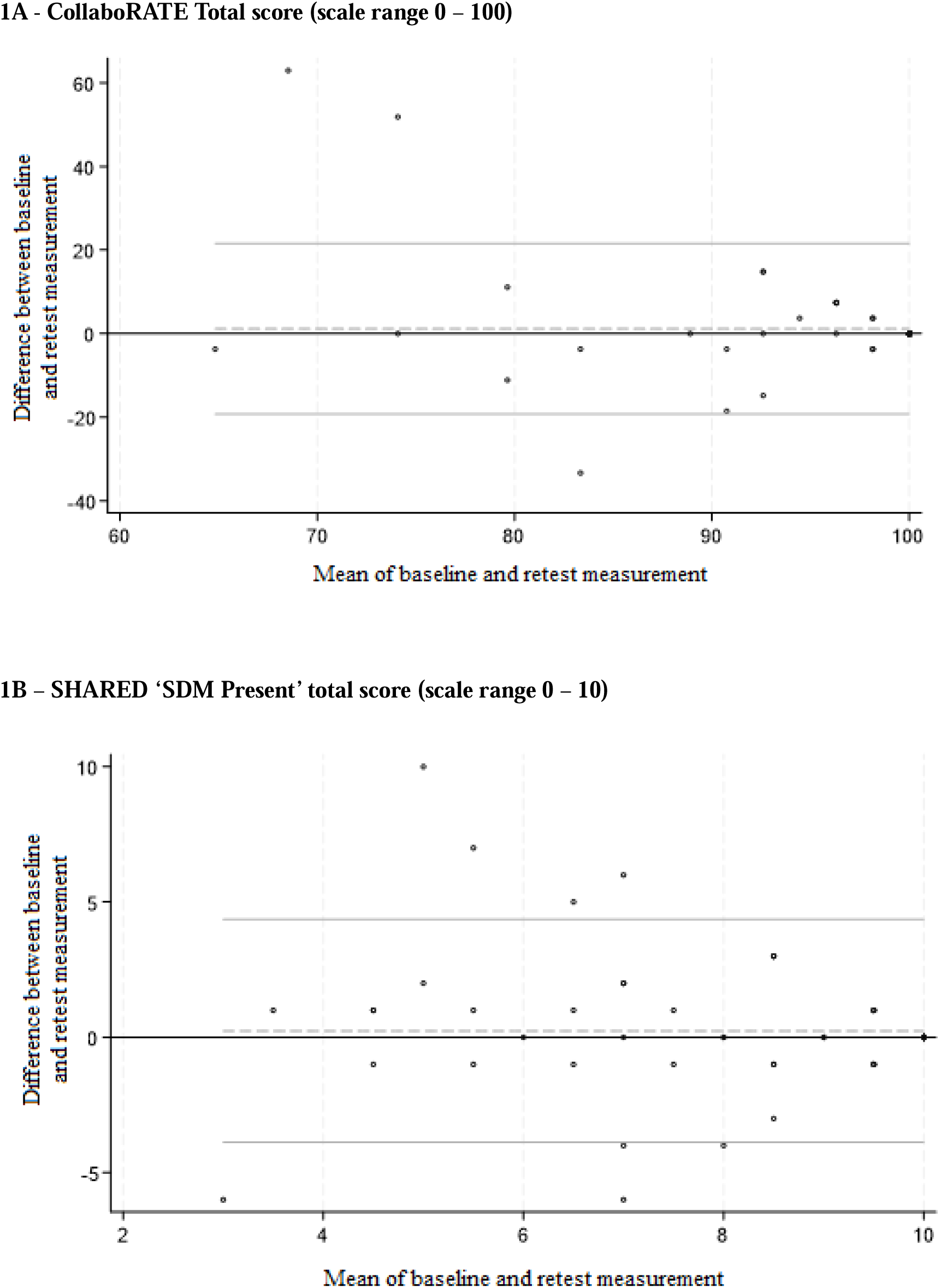
Bland-Altman plots demonstrating strength of agreement for each instrument between timepoints. Dashed line = mean difference. Solid line = 95% LoA (i.e. 95% of data points expected to be within these lines).

### Exploration of factors affecting perceived SDM stability

A total of nine patients were interviewed between January and March 2024, with interviews lasting an average of 30 minutes (range 13 - 65 minutes). Participant characteristics are summarised in **Table 2**. Two major themes were identified with subthemes that described factors affecting perceived SDM stability in the test-retest interval. These included: i) patients describing extended reasoning about the SDM process after their consultation and ii) a need for more support for SDM. Identified themes, subthemes and supporting quotes are illustrated in **Table 5**.

**Table 5.** Themes, sub-themes and supporting quotations identified from exploring the potential drivers for instability and score changes in the test-retest interval.

| Themes and subthemes |  |  | Supporting quotations. |
| --- | --- | --- | --- |
| i) | Patients described extended reasoning about the SDM process after their consultation | Patients reflected further on the consultation | *'[The period after the consultation/before surgery] is time to reflect on the whole thing, on the consultation, what information you were given in verbal and in the leaflets.'* - NBT051<br>*'a few days later I could think about it more and reflect about [the decision to have surgery] more...in some cases it might be you've come straight out [and] actually you've got so many things going around in your mind, what you tend to do is you drag out the bits that either thrill you, or make you nervous, and you focus on that. What [time] would do is give me a few days longer to think, instead of putting unsure because I couldn't remember' – NBT057* |
|  |  | Patients sought additional information from outside the consultation | *'I suppose I did do a lot of Googling and I know they say you should never Google medical things... I went in as much depth as I felt I could, and I suppose I sort of convinced myself that, yes, this probably is the way to go.'* - NBT067 |
|  |  | Repeat measurement of perceived SDM prompted reasoning | *'You [the study team] contacted me the second time, actually gave me a few more doubts because I thought, oh crikey, was I being a bit harsh in my answers? So, in a way, it did make me think more about the consultation and the doubts [with proceeding to surgery].' – NBT067*<br>*'I think straight after [the initial consultation] your mind is going in all sorts of directions, I think to do it [the initial survey] immediately, it might not get a very good result until it's sort of settled in a little bit and then you would probably get a clearer picture.'* - NBT051 |
| ii) | A need for more support for SDM | Patients needed time to process information | *'I could have said that I needed her to give it more time and sit down and go through it all again...at that point your head is swimming, and you can't think of anything. You think you've accepted [the decision to have surgery] when you walk home but I probably truly didn't accept it until a couple of weeks – well, a week later probably.'* – NBT048 |
|  |  | Unanswered questions led to ongoing reflection | *'Between doing the two surveys I've obviously stewed on it [the decision to have surgery] a little bit and wished that I knew the answers to those questions really because it does bother me...there's all these other things to consider [concerns around co-morbidities] that were a factor in making the decision, but it was never really discussed. It's just this extra concern that I need to know some answers to really.'* – NBT040 |
|  |  | Feeling led into a decision to have surgery | *'I wasn't given the option of 'think about it [surgery]' or 'do you want us to do it [surgery]?'. Nobody actually asked me that. She said, 'No, you haven't [got proven cancer] but we're going to remove it anyway... We just don't know but we want it [the possible cancer] out. I want to get you in and get it out straightaway.'.'* – NBT071 |
|  |  | Being unprepared for discussions around surgery | *'I wasn't expecting to actually be making a specific decision for myself. I think I was probably expecting to be talked through it [surgery] a little more... that made me feel as if I – I lacked confidence that I'd made the right decision there and then.'* – NBT067 |

#### Extended reasoning about the SDM process after the consultation

Patients have evolving thoughts about their experience of SDM in the days following the initial consultation where surgery is booked. Reflection is a fundamental part of the decision-making process, and patients are likely to develop new thoughts from their experience of SDM with the introduction of external sources of information, or by prompting through the process of retest measurement itself.

### Reflecting on the consultation

All participants stated that they reflected on their decision for surgery in the test-retest interval. They noted that the decision to have surgery is not something that can necessarily take place within the consultation itself and that a period of time is necessary to process the decision and their perceptions of SDM.

Reflection was common irrespective of the patient’s initial perceptions of SDM quality or certainty with their decision for surgery. For those who had lower perceived levels of SDM initially, the retest interval gave time to process information, to explore what was important to them and to consider the impacts that surgery would have for them.

- *‘Then as your life goes on for the next few days, you kind of think, “Oh, how’s it going to impact me?” …Then you kind of start to really think about it and it takes a while to actually accept it.’ – NBT048*

Patients who reported higher levels of SDM initially still reflected on their decision for surgery and on the SDM process, being able to re-affirm their decisions.

- *‘I must have been thinking, mulling it over in my mind and thinking well, yeah, I am more reassured than not reassured. It gave me more confidence…it made me more at peace with what was happening to me’ – NBT051*

### Patients sought additional information from outside the consultation

Patients reported receiving or seeking out sources of information from various sources (friends and family, hospital-provided information (leaflets, clinic letters), or online (websites, patient support forums)). Additional sources helped patients feel they had a more comprehensive understanding of the processes/risks/benefits of having surgery and had considered more opinions and options around the decision to have surgery. The gravity of surgical decisions led patients to perceive their involvement in the decision-making process as incomplete until additional information has been consulted. Processing any new information prompted further reflection and influenced their certainty about decision whether or not to have surgery, which may have ultimately affected their retest scoring.

- *‘I thought in this time that I’m waiting I’ve been really considering it and thinking about it and spoken to other people and got in touch with somebody on a Facebook group to actually find out what the alternatives are…Friends and relatives will ask some questions, so they’ll say oh did the surgeon explain what to do? And then it gives you time to think oh yeah, actually he did, or no, they didn’t say that.’ – NBT057*

### Repeat measurement of perceived SDM prompted reasoning

Patients reported that the survey content and contact with the study team prompted thoughts about their involvement in the decision to have surgery and had encouraged them to reflect on their consultation, which they may have not considered in as much detail otherwise.

- *‘‘[I’m] slightly prompted by answering the questions and going well, what did happen? I looked back on the information and what had been discussed in the appointment and thought well the discussion went a certain way but was I really making a choice? …In the context of the questionnaires and you subsequently asking me, it’s kind of like there were already decisions or recommendations already set out.’ - NBT039*

Patients commented that it was not only the understanding of the SDM process obtained by speaking to the study team that had an impact on their reflections, but also the timing of the second survey. Seven patients felt that the retest timepoint (day 5 – 10) was their preferred time for measurement to allow for information to be processed and reflected upon and therefore represented a truer reflection of their perceived involvement in SDM.

- *‘It was the second survey that made me think and want to reflect a bit more on it because you do the second one and…it just gives you time to sit and think about how I actually felt when I came away because you’re given all the information.’ – NBT048*

#### A need for more support for SDM

Participants reported barriers to their involvement in decision-making during their initial consultation which affected their decision-making capabilities for surgery. These barriers included overload of information, unanswered questions, or being led towards a decision by the surgeon. Patients demonstrated a desire to have some knowledge of the diagnosis or surgery before the consultation (often not received), which may have improved stability in the retest period.

### Patients needed time to process information

Patients expressed various reasons for not being able to process the information given to them at time of consultation. These included the volume and complexity of information, the ‘shock’ of the diagnosis being delivered, and the lack of time during consultation to have information repeated or reviewed.

- *‘He did explain what they’d do, but all this goes in but doesn’t… a lot of that I just hadn’t taken in.’ - NBT057*

### Unanswered questions led to greater reflection

Most patients were given the chance to ask questions in the consultation. Questions most often related to the surgical procedure itself (risks, benefits, alternatives), the peri-operative timeline, whether co-morbidities would affect the outcome of surgery and logistical queries. Due to a lack of time in the consultation or due to subsequent reflection on the SDM process, further questions arose in the test-retest interval after the decision for surgery had been made. Unanswered questions led to patients feeling less involved in the SDM process at retest measurement than immediately after the consultation. For some, these uncertainties led to a desire for further consultation with a surgeon to discuss their decision.

- *‘There’s just bits of hindsight where I’m going, oh should I have asked more, should I have explored this. One bit of what I want to explore more specifically about the risks and future implications if there had been a chance for another dialogue, I might have explored some of those areas.’ – NBT039*
- *‘The way I’m thinking about it now [is] ‘oh why didn’t I ask them more at the time?’. I didn’t come out of the appointment thinking ‘oh they didn’t answer my questions’ either.’- NBT039*

### Feeling led into a decision to have surgery

Patients described feeling that a decision had already been made by the HCP and this impacted on the extent to which they felt decision-making was truly shared. This was particularly the case for those booked for procedures to treat possible malignancy, where the alternatives to surgery were more likely (in the eyes of the HCP) to have poorer outcomes. At retest measurement, patients therefore felt that they may not have truly been presented a choice to make.

- *‘I looked back on the information and what had been discussed in the appointment and thought well the discussion went a certain way but was I really making a choice? …they wouldn’t do an operation if it wasn’t deemed necessary or appropriate, I’m just kind of reflecting on how much personal choice did I have in it. You know, were there any alternatives that could have been presented?’ – NBT039*
- *‘When I went back and read it [the retest survey] properly I was like oh, we didn’t really talk about other options…I’ve changed my opinion on the thing [survey] because there is no other option really.’ – NBT059*

### Being unprepared for discussions around surgery

Patients reported not feeling ready to discuss surgery as an option, largely due to the shock of diagnosis and complexities of surgical treatment. A lack of preparedness meant they did not know enough to formulate questions or have a meaningful discussion with the surgeon at the appointment. Patients expressed uncertainty with their decision for surgery, sought out other sources of information and had unanswered questions – these factors drove instability in the retest interval and led to lower scores at retest measurement.

- *‘I did feel a little bit thrown into the deep end. I think may be a more gentle [approach] or if I’d been prewarned… I just felt like, oh crikey, do I know enough to be making the right decision? …I should have gone in really with a list of questions so that you’re prepared, and you think right, these are all the questions I need to ask’’ - NBT067*

For patients that did feel prepared to discuss surgery (due to prior consultation or seeking out their own information), this instability was not present to the same extent.

- *‘Before I’d gone in, I’d done the research online myself. [The surgeon] just confirmed it and again gave the choice and gave me the information. She asked me what I thought, and I did say I’d need to go away and have a read but in my head I’d already made my mind up.’ – NBT048*
- *‘The nurse had said you come in and they just sort of talk you through the results and so on and, subject to the discussions, they would book you in for surgery…so I already knew in my mind that’s sort of what the plan was.’ - NBT039*

## DISCUSSION

### Principal findings

CollaboRATE and SHARED measurement instruments demonstrated fair-to-moderate agreement between initial and retest measurement with an ICC of 0.34 and 0.52 for CollaboRATE and SHARED total scores respectively. Analysis of agreement indicated no evidence of systematic bias, though it demonstrated that patients with lower initial scores may experience greater score changes on retest. Using qualitative methods to understand test-retest scores in the context of SDM is valuable to appreciate observed findings. Patients described how reasoning about decision making carried on for some time after their consultation and initial measurement. Some patients sought supplementary information from outside of the consultation or were prompted to reflect on decision making when asked to complete the retest measure. There was evidence to suggest those with lower initial scores and greater instability between measurement needed time to assimilate large amounts of information, address gaps in understanding, and think about what the consultation, and decision, meant for them.

This study highlights that considering when, how, and why SDM is measured is key to finding evidence that is meaningful to identify patient/professional needs to support SDM and develop clinical pathways. We found SDM in the surgical pathway to be perceived as a) a dynamic process extending beyond a single consultation or timepoint, and b) requiring proactive reasoning and problem solving from patients to engage further with their healthcare [15,31,32]. SDM measurement instruments, then, should not be seen as behaving in the same way as other SRQ measuring patient-outcome data, such as health-related quality of life scores, or symptom scores [33]. Stability of perceived SDM up to the point of surgery cannot be expected when the experience of care is likely to be impacted by proactive reasoning and further engagement with the clinical team along the pathway. Further, it seems likely that measuring the patient experience of SDM in a timely fashion with clinical teams could enhance the SDM process for patients during their preparation for surgery.

### In context of existing literature

The results of this study provide significant value in adding to the overall literature and level of evidence around SDM measurement instruments. In particular, the psychometric properties described (test-retest reliability, SEM and MDC) are rarely evaluated by existing studies [11]. In the limited studies in which test-retest reliability has been described in the context of SDM measurement, methodology of validation studies is poor, with few exploring stability. The findings from our mixed-methods study illustrate the difficulty of demonstrating psychometric rigour for instruments measuring cognitive processes. Patient perceptions of SDM and reflections about experience of healthcare are likely to be labile between test and retest points in the research process. They are likely to remain fluid until after a medical procedure has taken place, and possibly even thereafter [11].

Previous evaluation of CollaboRATE included intra-rater reliability assessed using simulated patient encounters (back pain) and a retest interval of 7 – 14 days. This demonstrated moderate strength of agreement using the ‘top score’ (ICC = 0.56), and excellent agreement using the ‘total score’ (ICC = 0.86). This contrasts with our study where we report moderate agreement for ‘top score’ (ICC = 0.47) and fair agreement for ‘total score’ (ICC = 0.34). Reasons for this disparity may include methodological differences in i) the use of simulated encounters and used intra-rater reliability rather than surgical patients making actual decisions and test-retest reliability, ii) a greater retest interval of 7-14 days rather than 5-10 days, and iii) the decision context of medical treatment for back pain rather than surgery). This is the first study to provide psychometric evidence for test-retest reliability, SEM and MDC for the SHARED questionnaire.

### Strengths and limitations

This study benefitted from a large sample size of patients for quantitative testing, beyond the minimum (50 participants) recommended for this type of psychometric testing. This study used real-world clinical encounters across seven different surgical specialties, and decisions concerned procedures of varying degrees of invasiveness, improving the generalisability of results. Study methodology was designed in accordance with the COSMIN and COREQ guidance, ensuring also that reporting standards were met to a high degree. All participants completed retest measurement within the pre-determined timeframe and there was no missing data. Novel to this study was the use of qualitative analysis of lived experience and stability in the retest interval to explore participants views which aided the interpretation of instrument scores. Interviews were completed alongside quantitative data collection, minimising potential recall bias and enabling emerging concepts to be explored in subsequent interviews.

There are several limitations. Data collection at both timepoints was performed digitally, which may exclude population groups with lower digital literacy or no access to the internet. While efforts were made to ensure that the wording of the CollaboRATE and the SHARED questionnaire items were the same, the survey platform, invitation wording and method of invitation were different between baseline and retest measurement timepoints due to resources not allowing for integration of retest measurement into the QI programme this study was embedded within. Available resources did not allow non-English language completion of SRQ or for interviews in this study. Despite efforts to recruit a diverse range of participants of various ethnicities, the sample lacked socio-demographic diversity (76% white, 62% female) and was recruited from a single NHS trust. The sample characteristics were reflective of the wider population served by the NHS trust, but there is a need for improved purposive sampling and to explore reasoning about SDM and engagement with healthcare from different ethnic and cultural group to ensure services can innovate and engage all patients in their healthcare [34,35].

### Implications for clinical and research practices

Both the CollaboRATE and SHARED measurement instruments have been used within a QI programme that sought to improve understanding of patient experiences of SDM through routine real-time measurement of SDM integrated into clinical pathways [8]. Results of psychometric testing and qualitative stability evaluation provide reliability data to inform how measures can be used and interpreted. Findings of this study indicate that patients focus on their reasoning about the decision and highlight their needs to support SDM along the surgical pathway.

Routine measurement of perceived SDM helps services understand what level of care patients feel they received when making healthcare decisions. It seems likely it may help identify further points in the service where patients needing additional engagement from the clinical team can talk through their choice of surgery, or other options. Repeat measurement up to the point of surgery may prompt and enable patients to deliberate and reflect on the decision-making process. This must be balanced with not introducing recall bias and to allow for enough time to carry out interventions to improve SDM. There is a need to better understand when the best time to measure SDM is in different settings and conditions, and how the measurement can be linked to a specific decision point or clinical encounter whilst capturing the complex and distributed nature of decision-making [15,16]. This could provide services with the rationale for employing timely and targeted interventions to improve the quality of the SDM process [2,3,8,36]. Clinicians measuring SDM as part of their practice should consider measurement timepoint(s) at the outset of pathway development with these issues in mind.

Clinicians, researchers and other stakeholders should think critically about what their SDM instruments are measuring, and why the healthcare context may impact on psychometric evaluations such as reliability. The dynamic nature and process of SDM make the measurement of patient-reported experiences of SDM complex and could limit the appropriateness of reliability as a psychometric property to consider when choosing an instrument and interpreting scores. This may differ to other widely used patient-reported outcome/experience measures (such as pain or quality of life scores) where the underlying constructs are less dynamic/static.

Though our results indicate a poor-to-moderate agreement between baseline and retest measurement in our patient cohort, this study did not explore what meaning can be inferred from quantitative scores, or what change in scores a patient perceives as important (the ‘minimal important change’).

This was outside of the scope of work for this study but remains an important area for further research to aid interpretability of SDM instrument scores.

## CONCLUSION

CollaboRATE and SHARED SDM measurement instruments demonstrate fair-to-moderate agreement between baseline and retest measures. Qualitative exploration revealed that patients continue to reason and reflect on SDM beyond the initial consultation, and that they have additional needs to support SDM during the initial consultation that can drive instability in the days following decision-making and implementation of (agreed) choice. Ongoing work is needed to determine the optimum time to measure SDM in surgical patients, to determine what change in scores is clinically important to patients or clinicians, and how best to prepare patients for surgical decision-making.

## Data Availability

All data produced in the present study are available upon reasonable request to the authors.

## ABBREVIATIONS

COREQ: COnsolidated criteria for REporting Qualitative studies
COSMIN: COnsensus-based Standards for selection of health status Measurement Instruments
HCP: Healthcare Professional
ICC: Intra-class Correlation Coefficient
LoA: Limits of Agreement
MDC: Minimal Detectable Change
NHS: National Health Service
NICE: National Institute for Health and Care Excellence
SRQ: Self-Reported Questionnaire
SEM: Standard Error of Measurement
SDM: Shared Decision-Making
UK: United Kingdom

## STATEMENTS AND DISCLOSURES

### Research ethics and consent

Approval for baseline data collection was completed as part of a quality improvement proposal at North Bristol NHS Trust (Reference Q80008). Approval for research using this baseline data was granted by the NHS Health Research Authority North West – Liverpool Central Research Ethics Committee (Reference 21/PR/0345). Ethics approval for research activities including re-test measurement and qualitative interviews was granted by the NHS Health Research Authority North West – Liverpool Central Research Ethics Committee (Reference 21/NW/0091). Participants provided electronic consent through a link to a secure online survey platform (JISC) prior to commencing study activity.

### Data availability

Data are available on reasonable request. The deidentified datasets used and/or analysed during the current study are available from the corresponding author on reasonable request.

### Licenses and permissions

An academic/research license was obtained for use of The SHARED Questionnaire in this study from the University of Leeds (Licence Order 18LC3K). The SHARED Questionnaire can be obtained at https://licensing.leeds.ac.uk/product/shared-questionnaire.

The CollaboRATE measurement instrument is freely available for use under a Creative Commons Attribution-NonCommercial-NoDerivs 3.0 License. Permissions to use the CollaboRATE questionnaire for research purposes and reproduce the questionnaire for publication were obtained via registration of this study with the instrument developers. The CollaboRATE questionnaire can be obtained at https://www.glynelwyn.com/collaborate.html.

### CRediT author statement

**V.G.:** Methodology, Validation, Formal analysis, Investigation, Data Curation, Writing - Original Draft, Writing - Review & Editing, Visualisation, Project administration; **C.H.:** Conceptualisation, Methodology, Data Curation, Writing - Review & Editing, Supervision, Project administration; **H.L.B.:** Validation, Resources, Writing - Review & Editing; **S.L.:** Validation, Formal analysis, Writing - Review & Editing; **A.J.:** Methodology; **D.H.:** Resources, Data Curation; **J.B.:** Conceptualisation, Supervision, Funding acquisition; **K.A.:** Conceptualisation, Writing – Review & Editing; **A.G.K.M:** Conceptualisation, Methodology, Formal analysis, Writing - Review & Editing, Supervision, Funding acquisition

All authors have read and approved the final version of the manuscript and agreed with the order of presentation of the authors

