## Appendix 1 for "Reliability testing of patient-reported measurement instruments of Shared Decision-Making within surgical treatment pathways: a mixed-methods study"

**-Template Topic Guide-**

| Introduction (5mins) | - Researcher introduction to interviewee (name, role, inability to answer care-specific questions), offer thanks   *‘Thank you for taking the time to talk to me about your experiences with decision making at Southmead Hospital. As part of the project, we are speaking to patients to try and support improvements to patients experience with SDM. It is really valuable for us to understand your personal experience so we can improve things for future patients.’*   - Reminder of anonymity, confidentiality and that participants can withdraw at any time - Check if any questions from interviewee - Check if OK to audio-record --> **Switch on recorder**   *‘Before we start, do you have any questions about the study or the information that were sent to you? Can I also check again that are you happy for our conversation to be recorded?’* | |
| --- | --- | --- |
| Warm up and understanding of SDM (5mins) | - Explain what SDM is - Explain what reliability testing is - Brief explanation of wider context within quality improvement project - Reason for inviting participants (understand more about their responses, how the hospital can improve care) | |
| Exploring views on: | | |
| Initial consultation | - Explore what their knowledge of the surgery they are booked for involves, what the conversation with the consultant involved. | |
| Initial test scores (5mins) | - *Could you tell me a bit more about how you responded to the initial survey?* - *What were your initial thoughts when receiving the survey?* - *Why did you answer question X in the way that you did?* - *Were there any factors outside of the consultation that affected how you responded to the survey?* | - Read out questions from survey if required. - *Why did you score ‘question X’ that way?* |
| Re-test scores (10mins) | - Explore specific differences e.g. score increase or decreases - Explore similarities – why did your score not change? (in the context of new thoughts/views but no change in score) - *Could you tell me a bit more about how you responded to the second survey?* - *Can you explain why you thought/felt X/Y/Z?* | - Remind participants of survey responses and questions if required. - *I am interested in your responses to ‘question X’ - why do you think you responded in this way?* - *Why do you think your response differed between the first and second times you filled out this survey?* |
| Test-retest interval (10mins) | - If decrease in scores/poor baseline SDM -->   - *How can/could the surgical team have made things better?*   - *What would the surgical team have to do so that your scores go up?* - If increase in scores -->   - *What changed to make your experience of SDM better?* - *Were there any factors outside of the consultation that affected how you responded to the repeat survey?* - *What aspect of your experience of your initial appointment should be fed back to the surgical team? Why? How?* - *What aspect of your thoughts in the following days/second survey response should be fed back to the surgical team? Why? How?* - *How do you feel about going ahead with the planned surgery now versus when you left the initial appointment?* | - *Have you had any further thoughts about your surgery or consultation with a surgeon since completing the first survey?* - *Did completing the survey a second time make you think about your surgery or consultation in more detail/raise new thoughts?* - *Are there any questions in particular that made you think more about your surgery or consultation? Why? What thoughts/feelings did you have?* - *When do you think the best time for the surgical team to get feedback on your thoughts is?* |
| Wrap up (5mins) | - Explain next steps - Thank participant   *‘Thank you for sharing your experiences, that was really helpful for us to figure out how we improve SDM for patients in the future. Do you have any final thoughts about the survey or the project or any other questions?’* | |
